# No causal association between inguinal hernia and aortic aneurysm using Mendelian randomization analysis

**DOI:** 10.1101/2023.04.21.23288915

**Authors:** Sicheng Yao, Hongbo Ci

**Author notes:** Corresponding author: Hongbo Ci, MD, PhD, Address: 91 Tian Chi Road, Urumqi, 830001, People’s Republic of China.

## Abstract

Aortic aneurysm (AA) is a serious disease that affects the aging population worldwide. Potential risk factors such as inguinal hernia has been suggested by conventional studies could contribute to AA. The aim of our study was to clarify the causal association between inguinal hernia and AA using Mendelian randomization (MR) analysis. Summary statistics data for the associations of inguinal hernia were derived from a recently published large genome-wide association study including 18,791 inguinal hernia cases and 93,955 controls in UK Biobank. Corresponding data of AA were extracted from FinnGen, comprising 7,603 cases and 317,899 controls. The causal association was assessed using MR-egger, weighted median, and inverse variance weighting methods, and compared to observational estimates previously published. Our analysis found no convincing causal effect between genetically predicted inguinal hernia and the risk of AA (odds ratio [OR] = 1.05, 95% confidence interval [CI] = 0.85–1.31, *p* = 0.65), AAA (OR = 1.15, 95% CI = 0.92–1.46, *p* = 0.22), and TAA (OR = 1.05, 95% CI = 0.85–1.30, *p* = 0.67). The results are in contrast to previous observational evidence suggesting a harmful effect of inguinal hernia.

## 1. Introduction

Aortic aneurysm (AA) is an abnormal enlargement of the wall of any segment aorta, based on the different location it can be classified as abdominal aortic aneurysm (AAA) or thoracic aortic aneurysm (TAA)[1]. It is a complex and deadly disease of the aging population, prevalence of AAA in men increases by 6% per decade after 65 years of age[2]. This disease often remains asymptomatic until it progresses to the point of rupture or dissection, making it difficult to diagnose and treat. Although medical advancements have made it easier to detect AA at an early stage, there are currently no medical therapies available to prevent its continuous dilation[3]. Surgery is currently the only valid treatment, with open surgery repair (OSR) and endovascular aneurysm repair (EVAR) being the most common methods[4]. While EVAR is less invasive and results in fewer procedural deaths, it has unique complications such as endoleaks that can lead to greater morbidity and mortality[4].

Although TAA has different pathophysiological changes, AAA and TAA both share common risk factors such as smoking, age over 65, male sex, and family history[1,5,6]. Diseases like pulmonary emphysema and inguinal hernia have been found possible contributions to AAA[7]. Inguinal hernia is defined as weakness in the transversalis fascia and shares a common mechanism with AAA that involves degeneration of the connective tissue mediated by matrix metalloproteinases[8,9]. Moreover, AA and inguinal hernia both increased risks among first-degree relatives of people suggesting that there are likely identifiable genetic risk factors[7]. According to a recently published large retrospective cohort study (n = 67,732), geriatric patients with hernia had a higher incidence rate, and adjusted hazard ratio of AA, as well as a marginally higher aneurysm rupture rate than those without[7]. However, another cohort study (n = 18,331) focused only on male patients in Denmark reported no associations between inguinal hernia and AAA. Whether inguinal hernia has a causal inference on AA remains uncertain[10].

This inconsistent results from conventional observational studies can be explained by some methodological limitations, such as reverse causation and confounding bias[11,12]. Recent studies using the Mendelian randomization (MR) strategy selected genetic variations as instruments to identify causal relationships between outcomes and exposures[5,13]. Since genetic variations are assigned at random during conception, confounding and reverse causality can be substantially minimized and diminished, allowing a more accurate evaluation of causal relationships[14]. To date, only a small number of risk factors or candidate biomarkers have been investigated by MR in AA[15]. This study aims to use MR analysis to determine if there is a causal relationship between inguinal hernia and AA, which could provide important insights into the development and prevention of AA.

## 2. Materials and Methods

### 2.1 Study design

This study was designed using a two-sample Mendelian randomization method, followed by the Strengthening the Reporting of Observational Studies in Epidemiology using Mendelian Randomization (STROBE-MR) reporting guidelines[12]. Three fundamental presumptions (relevance, independence, and exclusion restriction) must be satisfied in this MR analysis[16]: (1) Instrument variables (IVs) are strongly associated with the inguinal hernia; (2) IVs are not associated with confounders of the exposure-outcome relationship; (3) IVs have an effect on the AA only through the inguinal hernia.

An overview of this study design is depicted in Figure. 1. Ethics approval is not required for this study as all summary statistics were extracted from publicly available biobanks and had already been approved by their local ethical committees.

**Figure 1.**
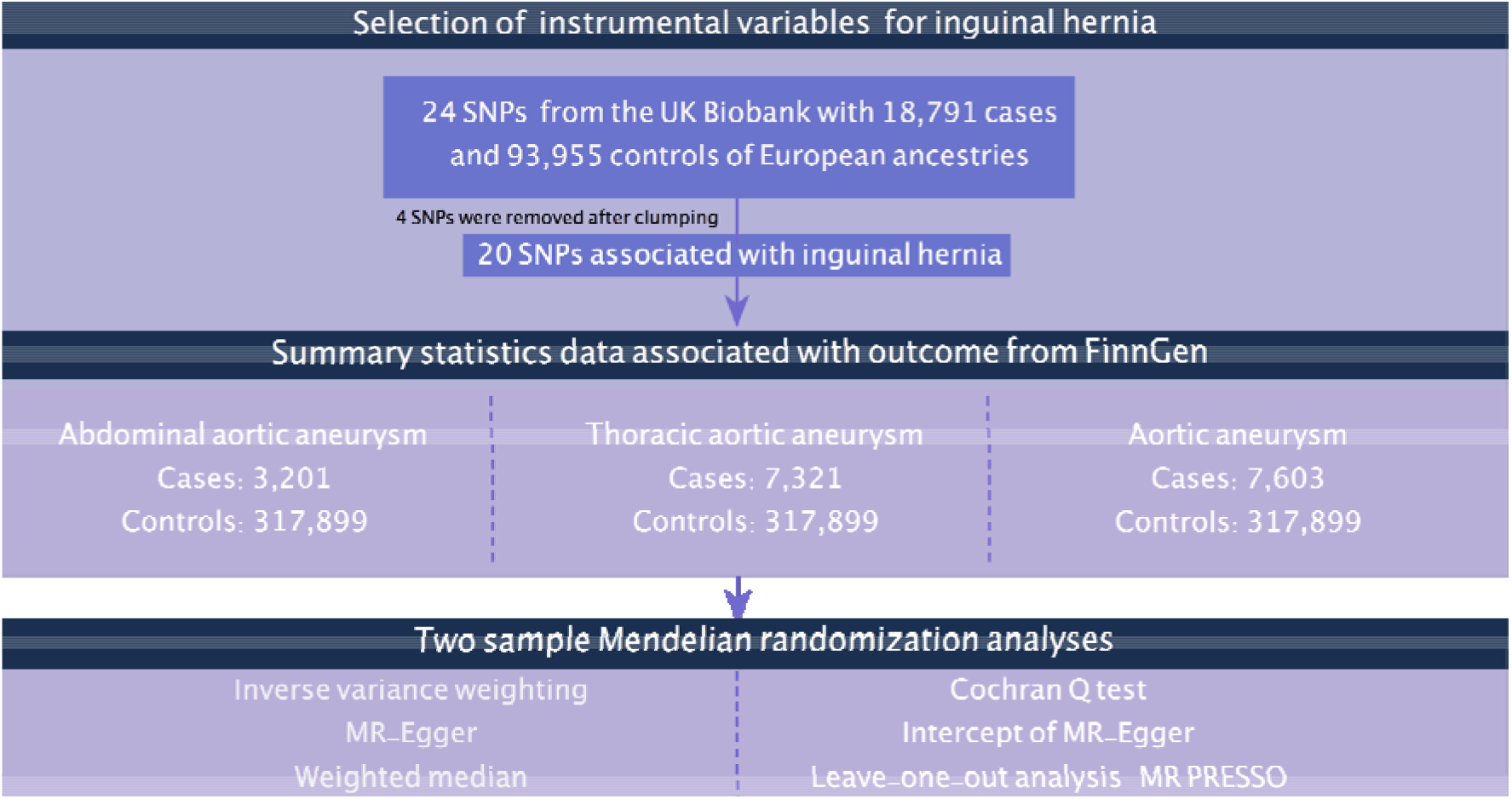
Study overview of this mendelian randomization.

### 2.2. Genetic instrument

Summary statistics data for the associations of single nucleotide polymorphisms (SNPs) with inguinal hernia were derived from a recently published large genome-wide association study (GWAS) including 18,791 inguinal hernia cases and 93,955 controls in UK Biobank[17]. Cases were defined as those who had diagnostic and/or operative coding for only inguinal hernia. The study further performed SNP quality control based on deviations from Hardy-Weinberg equilibrium (*p* < 10^−4^), minor allele frequency (MAF) <0.01, as well as on visual inspection of autosomal heterozygosity against call rate[17]. Twenty-eight SNPs associated with inguinal hernia were selected as IVs. The linkage disequilibrium (LD) across 24 SNPs was estimated using the clump function in the TwoSampleMR package (set as r^2^ < 0.001 and clump window > 10,000 kb) based on the European ancestry LD reference panel from the 1000 Genomes Project. After removed 4 variants due to LD with other variants or absence from the LD reference panel, all of the remaining 20 SNPs were available in the outcome summary data. The gene EFEMP1 (rs56046099, rs1899888) encodes fibulin-3 was the most highly expressed of the inguinal hernia risk loci in mouse connective tissue, several studies also identified EFEMP1 interactions in inguinal hernia pathophysiology, confirming the validity of the selected genetic variants[8,18].

### 2.3. Outcome data

Corresponding data for the associations of AA were obtained from the FinnGen consortium (R8 release)[19]. The FinnGen consortium included 342,499 Finnish ancestry participants, non-Finnish origin were not included and had been adjusted for age, sex and genotyping batch[19]. We have a final summary statistics data of AA comprising 7,603 cases and 317,899 controls in Finnish ancestry. Subtypes were also included in this study since the FinnGen consortium contained the endpoints of AAA and TAA. The FinnGen study’s endpoints were constructed from the register codes using the Finnish version of the International Classification of Diseases[19]. There was no sample overlap between exposure and outcome summary statistics data.

### 2.4. Statistical analysis

All statistical analyses were conducted using R software v4.2.0 (The R Foundation for Statistical Computing, Vienna, Austria). The TwoSample MR package for R was utilized for all MR analyses. An F-statistic was computed for each SNP to measure instrument strength and assess for weak instrument bias[20]. Primary MR analyses included MR-Egger, weighted median, and inverse variance weighting (IVW)[21,22]. For IVW as the main analysis, we used a random effect model if there was heterogeneity existed or a fixed effect model will be applied if there was no heterogeneity. Estimates for each SNP were computed by dividing the beta coefficient of the SNP outcome estimate by the beta coefficient of the SNP exposure estimate, and consistent results were obtained among the three different methods to enhance reliability. Additional analyses were conducted to evaluate possible heterogeneity and pleiotropy, including the MR-Egger intercept and Cochran’s Q test[12]. MR-Pleiotropy Residual Sum and Outlier methods (MR-PRESSO) were also used to assess and correct horizontal pleiotropy[23]. Leave-one-out analysis was conducted to assess the association between genetic instruments and exposure reliability.

## 3. RESULTS

In total, we incorporated 20 SNPs in this MR analysis, detailed combined SNPs information is listed in Table S1. F-statistics for all SNPs used as instrumental variables were greater than 10, indicating a low likelihood of weak instrumental variable bias. No convincing casual effect between genetically predicted inguinal hernia and risks of AA (odds ratio [OR] = 1.05, 95% confidence interval [CI] = 0.85–1.31, *p* = 0.65), AAA (OR = 1.15, 95% CI = 0.92–1.46, *p* = 0.22), and TAA (OR = 1.05, 95% CI = 0.85–1.30, *p* = 0.67) according to the results of IVW analysis. The MR-Egger and weighted median models, which are more robust to directional pleiotropy, produced similar results which make our results more reliable (Figure.2). The MR-Egger intercepts suggested that no evidence for significant directional pleiotropy. There is no heterogeneity in the outcome of AAA and TAA by Cochran’s Q statistical analysis, the outcome of AA showed substantial heterogeneity but no distinct outliers were detected. Leave-one-out analysis suggested that the associations observed in our study were unlikely to be influenced by certain extreme SNPs, further confirmed the robustness of the findings (Figure S1-S3).

**Figure 2.**
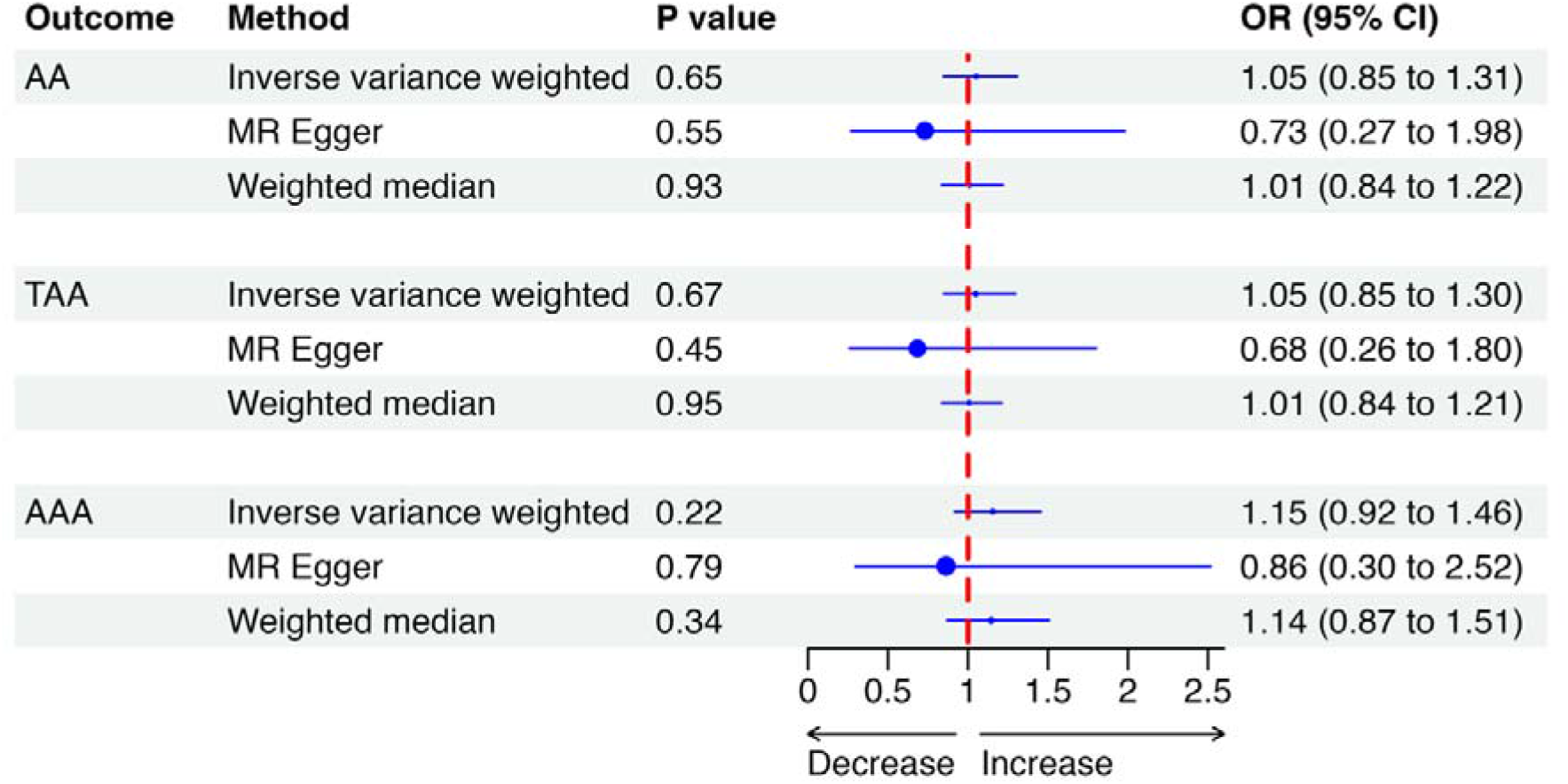
Results of three different mendelian randomization methods to investigate genetic association between inguinal hernia and aortic aneurysms. Abbreviations: MR, mendelian randomization; AA, aortic aneurysm; TAA, thoracic aortic aneurysm; AAA, abdominal aortic aneurysm; CI, confidence interval; OR, odds ratio.

## 4. DISCUSSION

To the best of our knowledge, this is the first MR study to investigate the causal effects between inguinal hernia and AA, as well as its subtypes. The IVW analysis found no casual relationships between inguinal hernia and the development of aortic aneurysms. In addition, the results of our two complementary analyses (the MR-Egger and weighted median models) also reached the same conclusion.

Previous study by Cannon and colleagues in 1984 suggested a possible link between inguinal hernia and aortic aneurysm, which was then supported by many others[7,24–26]. However, the results of conventional observational studies have been inconsistent. The majority of studies mainly focused on AAA, with a maximum of 600 cases. One study used data from an applied questionnaire and found a relatively higher prevalence of AAA in inguinal hernia patients (27% vs. 31%) than those without[7]. The study’s logistic regression analysis showed that the adjusted odds ratio was 1.2 (95%CI = 1.0–1.4) after controlling for other risk factors for AAA. Another cohort study enrolled 18,331 men aged 65-76 years from the Danish National Patients Registry, with a median of 34.9 years of follow-up. However, no significant association was found between inguinal hernia and AAA (OR = 0.86, 95% CI = 0.68-1.09; adjusted OR = 0.94, 95%CI = 0.75-1.20)[10]. Both studies were unable to eliminate the possibility that some individuals might have had undiagnosed AAA prior to receiving a hernia diagnosis. Therefore, the prevalence of AAA in inguinal hernia patients might be overestimated or underestimated.

In addition to the European ancestry studies mentioned above, a recent large-scale prospective cohort study conducted on the Asian population reveal that patients with hernia had a greater incidence rate and hazard ratio of aortic aneurysm compared to those without hernia (adjusted subdistribution hazard ratio [sdHR] = 1.34, 95% CI = 1.02-1.76)[27]. And geriatric patients over the age of 65 had a particularly higher hazard ratio (sdHR = 1.44, 95% CI = 1.07-1.94). The study also reports a marginally greater risk of TAA (sdHR = 1.66, 95% CI = 0.96-2.86) or AAA (sdHR = 1.36, 95% CI = 0.96-1.94) rupture among individuals with hernia[27]. Similar to previous research, the natural course of the aneurysm and its potential correlation in these studies were unable to determine due to the fact that the time of aneurysm development was not reflected by the time of its diagnosis. On the other hand, individuals who have hernia may undergo imaging examinations more frequently. While the frequencies of various imaging techniques have been tallied by the authors, it cannot be dismissed that individuals with hernia might receive a more extensive evaluation at the same scan frequency.

As most studies cannot be used to draw causal inferences, we therefore conducted this MR study to further investigate this issue. Different from the methodological design and larger sample sizes of the outcome datasets, our MR estimates had high precision, as indicated by relatively narrow CIs. The OR obtained through MR yielded similar results to previous observational studies. However, no causal link between inguinal hernia and AA has been found, a deviation from prior studies. The complexity of the pathogenesis of aneurysm formation and limited knowledge may lead to discrepancies in this research findings. Several studies have found a 20% lower risk of AAA in individuals with diabetes compared to those without. However, a recent two-stage MR study revealed that genetic predisposition to diabetes did not confer protection against AAA. It is possible that previous findings were biased by metformin, a common diabetes medication[15].

Various physiological pathways, including inflammation, lipid metabolism, extracellular matrix (ECM) remodeling, and angiogenesis, have been found to contribute to AAA formation, although the pathogenesis of TAA is different, the discussion in this paper is based on the mechanisms shared by both and is mainly focused on AAA [2]. It is important to note that dysregulation of connective tissue plays a significant role both in the development of inguinal hernia and AAA. The maintenance of connective tissue depends on the physical and biochemical properties of the ECM, mainly composed of collagen and elastin proteins. And an appropriate balance of matrix metalloproteinases (MMPs) and tissue inhibitors of metalloproteinases (TIMPs) is essential to stable ECM homeostasis[2,28]. Findings reveal that collagen levels are significantly lower in transversalis fascia samples derived from patients with inguinal hernia compared to controls. And the ratio of MMP to TIMP expression is higher in both inguinal hernia and AA, leading to ECM breakdown[26,28]. These findings suggest that there may be risk factors impacting both hernia and aortic aneurysm through common pathways. Observational and MR studies have confirmed smoking and abdominal obesity as prevalent factors[6]. Smoking can increase elastin degradation and aneurysm size, supporting previous observational studies that could elucidate relationships through confounders such as smoking[29,30]. However, identifying individuals who are at a high risk of developing aneurysms and would benefit from screening programs remains a challenging task. Additional studies are needed to better understand the interplay between risk factors and their influence on the development of aneurysm, which can help improve the identification and management of high-risk individuals.

Our study has several strengths. Firstly, unlike previous observational studies, which utilized case-control or cross-sectional designs, our results from two-sample MR analyses are more precise and less susceptible to confounding bias and reverse causation. Furthermore, the consistency of the results from three different MR statistical models reinforces our estimates. Moreover, the inclusion of several sensitivity analyses enhances the robustness of our study results by accounting for possible confounders or biases in the data. Secondly, the summary statistics data used in our study were extracted from large-scale consortia and biobanks, which allowed for large sample sizes of AA compared to previous studies, with a maximum of 400 cases. Thirdly, only a few AA risk factors or biomarkers have been investigated using MR methods. Fourthly, the use of publicly available datasets created a more transparent research process by allowing other researchers to replicate our findings.

Although our study provides significant insights into the genetic assumption of aortic aneurysms, it is important to note several limitations. One of the primary limitations of our analysis is that it focused solely on European individuals. Therefore, the generalizability of our findings to populations with different ethnicities remains unclear. Second, the interpretation of MR estimates in the presence of heterogeneity should be cautious, although additional sensitivity analyses have been conducted to assess the robustness of the results. Thirdly, even though we utilized two separate data sources, the sample size of AA was limited, suggesting that further research is needed to carry out well-designed GWAS with larger sample sizes and uniform aneurysm definition to accurately determine the genetic associations of AA. Moreover, the lack of summary statistics data regarding aneurysm rupture or aortic diameter makes it difficult to deeper interpret the natural history of AA. Conducting additional research on genetic factors associated with aneurysm rupture or aortic diameter could be valuable in improving patient outcomes and developing more effective screening and treatment protocols for AA.

In summary, our MR analysis does not support a causal relationship between inguinal hernia and aortic aneurysm in European populations. These findings contrast with previous observational evidence suggesting a harmful effect of inguinal hernia. Therefore, further research is needed to better understand the interplay between risk factors and their impact on aneurysm development, which can help improve the identification and management of high-risk individuals.

## Supplementary Materials

Supplementary data to this article can be found online.

## Data Availability

All data used in the current study are publicly available GWAS summary data. The MR analysis code used in R can be found at https://mrcieu.github.io/TwoSampleMR/artic les/index.html.

https://mrcieu.github.io/TwoSampleMR/articles/index.html

## Authors’ contributions

Conceptualization: SCY.

Data curation: SCY.

Formal analysis: SCY, HBC.

Funding acquisition: HBC.

Investigation: SCY, HBC.

Methodology: SCY, HBC.

Project administration: HBC.

Software: SCY.

Supervision: HBC.

Validation: HBC.

Visualization: SCY.

Writing – original draft: SCY. Writing – review & editing: all authors.

## Acknowledgements

We want to acknowledge the participants and investigators of t the UK Biobank and FinnGen projects.

## Availability of data and materials

All data used in the current study are publicly available GWAS summary data. The MR analysis code used in R can be found at https://mrcieu.github.io/TwoSampleMR/articles/index.html.

## References

1. Mathur, A.; Mohan, V.;Ameta, D.;Gaurav, B.;Haranahalli, P. Aortic Aneurysm. Journal of Translational Internal Medicine 2016, 4, 35–41.

2. Quintana, R.A.;Taylor, W.R. Cellular Mechanisms of Aortic Aneurysm Formation. Circ Res 2019, 124, 607–618.

3. Bossone, E.;Eagle, K.A. Epidemiology and Management of Aortic Disease: Aortic Aneurysms and Acute Aortic Syndromes. Nat Rev Cardiol 2021, 18, 331–348.

4. Patel, R.;Sweeting, M.J.;Powell, J.T.;Greenhalgh, R.M. Endovascular versus Open Repair of Abdominal Aortic Aneurysm in 15-Years’ Follow-up of the UK Endovascular Aneurysm Repair Trial 1 (EVAR Trial 1): A Randomised Controlled Trial. The Lancet 2016, 388, 2366–2374.

5. Ibrahim, M.;Thanigaimani, S.;Singh, T.P.;Morris, D.;Golledge, J. Systematic Review and Meta-Analysis of Mendelian Randomisation Analyses of Abdominal Aortic Aneurysms. IJC Heart & Vasculature 2021, 35, 100836.

6. Zhou, J.;Lin, J.;Zheng, Y. Association of Cardiovascular Risk Factors and Lifestyle Behaviors with Aortic Aneurysm: A Mendelian Randomization Study. Front. Genet. 2022, 13, 925874.

7. Pleumeekers, H.J.C.M.;de Gruijl, A.;Hofman, A.;van Beek, A.J.;Hoes, A.W. Prevalence of Aortic Aneurysm in Men with a History of Inguinal Hernia Repair. British Journal of Surgery 2002, 86, 1155–1158.

8. Jorgenson, E.;Makki, N.;Shen, L.;Chen, D.C.;Tian, C.;Eckalbar, W.L.;Hinds, D.;Ahituv, N.;Avins, A. A Genome-Wide Association Study Identifies Four Novel Susceptibility Loci Underlying Inguinal Hernia. Nat Commun 2015, 6, 10130.

9. Fadista, J.;Skotte, L.;Karjalainen, J.;Abner, E.;Sørensen, E.;Ullum, H.;Werge, T.;iPSYCH Group; Werge, T.;Hougaard, D.M.;et al. Comprehensive Genome-Wide Association Study of Different Forms of Hernia Identifies More than 80 Associated Loci. Nat Commun 2022, 13, 3200.

10. Henriksen, N.A.;Sorensen, L.T.;Jorgensen, L.N.;Lindholt, J.S. Lack of Association between Inguinal Hernia and Abdominal Aortic Aneurysm in a Population-Based Male Cohort. British Journal of Surgery 2013, 100, 1478–1482.

11. Yuan, S.;Bruzelius, M.;Larsson, S.C. Causal Effect of Renal Function on Venous Thromboembolism: A Two-Sample Mendelian Randomization Investigation. J Thromb Thrombolysis 2022, 53, 43–50.

12. Skrivankova, V.W.;Richmond, R.C.;Woolf, B.A.R.;Davies, N.M.;Swanson, S.A.;VanderWeele, T.J.;Timpson, N.J.;Higgins, J.P.T.;Dimou, N.;Langenberg, C.;et al. Strengthening the Reporting of Observational Studies in Epidemiology Using Mendelian Randomisation (STROBE-MR): Explanation and Elaboration. BMJ 2021, n2233.

13. Yuan, S.;Bruzelius, M.;Xiong, Y.;Håkansson, N.;åkesson, A.;Larsson, S.C. Overall and Abdominal Obesity in Relation to Venous Thromboembolism. J. Thromb. Haemost. 2021, 19, 460–469.

14. Smith, G.D. Capitalizing on Mendelian Randomization to Assess the Effects of Treatments. 2007, 100.

15. Morris, D.R.;Jones, G.T.;Holmes, M.V.;Bown, M.J.;Bulbulia, R.;Singh, T.P.;Golledge, J. Genetic Predisposition to Diabetes and Abdominal Aortic Aneurysm: A Two Stage Mendelian Randomisation Study. European Journal of Vascular and Endovascular Surgery 2022, 63, 512–519.

16. Davies, N.M.;Holmes, M.V.;Davey Smith, G. Reading Mendelian Randomisation Studies: A Guide, Glossary, and Checklist for Clinicians. BMJ 2018, k601.

17. Ahmed, W.U.-R.;Patel, M.I.A.;Ng, M.;McVeigh, J.;Zondervan, K.;Wiberg, A.;Furniss, D. Shared Genetic Architecture of Hernias: A Genome-Wide Association Study with Multivariable Meta-Analysis of Multiple Hernia Phenotypes. PLoS ONE 2022, 17, e0272261.

18. Livingstone, I.;Uversky, V.N.;Furniss, D.;Wiberg, A. The Pathophysiological Significance of Fibulin-3. Biomolecules 2020, 10, 1294.

19. Kurki, M.I.;Karjalainen, J.;Palta, P.;Sipilä, T.P.;Kristiansson, K.;Donner, K.;Reeve, M.P.;Laivuori, H.;Aavikko, M.;Kaunisto, M.A.;et al. FinnGen: Unique Genetic Insights from Combining Isolated Population and National Health Register Data; Genetic and Genomic Medicine, 2022;

20. Burgess, S.;Thompson, S.G.;CRP CHD Genetics Collaboration Avoiding Bias from Weak Instruments in Mendelian Randomization Studies. International Journal of Epidemiology 2011, 40.

21. Bowden, J.;Davey Smith, G.;Haycock, P.C.;Burgess, S. Consistent Estimation in Mendelian Randomization with Some Invalid Instruments Using a Weighted Median Estimator. Genet. Epidemiol. 2016, 40, 304–314.

22. Bowden, J.;Davey Smith, G.;Burgess, S. Mendelian Randomization with Invalid Instruments: Effect Estimation and Bias Detection through Egger Regression. International Journal of Epidemiology 2015, 44, 512–525.

23. Verbanck, M.;Chen, C.-Y.;Neale, B.;Do, R. Detection of Widespread Horizontal Pleiotropy in Causal Relationships Inferred from Mendelian Randomization between Complex Traits and Diseases. Nat Genet 2018, 50, 693–698.

24. Cannon, D.J. Abdominal Aortic Aneurysm, Leriche’s Syndrome, Inguinal Herniation, and Smoking. Arch Surg 1984, 119, 387.

25. Golledge, J.;Reeve, T.;Norman, P.E. ABDOMINAL AORTIC ANEURYSM, INGUINAL HERNIAS AND EMPHYSEMA. ANZ Journal of Surgery 2008, 78, 1034–1034.

26. Hinterseher, I.;Miszczuk, M.;Corvinus, F.;Zimmermann, C.;Estrelinha, M.;Smelser, D.T.;Kuivaniemi, H. Do Hernias Contribute to Increased Severity of Aneurysmal Disease among Abdominal Aortic Aneurysm Patients? Aorta (Stamford) 2021, 09, 009–020.

27. Hung, K.-C.;Chang, Y.-J.;Sun, C.-K.;Wang, J.-J.;Chen, Y.-C.;Weng, S.-F.;Chu, C.-C. Association of Hernia with Subsequent Aortic Aneurysm in Geriatric Patients. The Journal of Thoracic and Cardiovascular Surgery 2021, 162, 1668–1677.e2.

28. Maguire, E.M.;Pearce, S.W.A.;Xiao, R.;Oo, A.Y.;Xiao, Q. Matrix Metalloproteinase in Abdominal Aortic Aneurysm and Aortic Dissection. Pharmaceuticals 2019, 12, 118.

29. Aune, D.;Schlesinger, S.;Norat, T.;Riboli, E. Tobacco Smoking and the Risk of Abdominal Aortic Aneurysm: A Systematic Review and Meta-Analysis of Prospective Studies. Sci Rep 2018, 8, 14786.

30. Bergoeing, M.P.;Arif, B.;Hackmann, A.E.;Ennis, T.L.;Thompson, R.W.;Curci, J.A. Cigarette Smoking Increases Aortic Dilatation without Affecting Matrix Metalloproteinase-9 and -12 Expression in a Modified Mouse Model of Aneurysm Formation. Journal of Vascular Surgery 2007, 45, 1217–1227.e2.

