## Supplementary material for "No causal association between inguinal hernia and aortic aneurysm using Mendelian randomization analysis": S1

**Supplement material 1**

S1 Table 1 Genetic variants selected as instrument variables in this study.

S1 Table 2 Full results of three different Mendelian randomization methods.

S1 Table 3 Pleiotropy test results.

S1 Table 4 Heterogeneity test results.

S1 Figure 1 MR leave-one-out sensitivity analysis for the effect of the inguinal hernia SNPs on AA.

S1 Figure 2 MR leave-one-out sensitivity analysis for the effect of the inguinal hernia SNPs on TAA.

S1 Figure 3 MR leave-one-out sensitivity analysis for the effect of the inguinal hernia SNPs on AAA.

**S1 Table 1 Genetic variants selected as instrument variables in this study.**

| chr | pos | SNP | effect_allele | other_allele | eaf | beta | pval | se |
|---|---|---|---|---|---|---|---|---|
| 1 | 9443340 | rs1106370 | A | G | 0.42 | 0.06765865 | 1.00E−08 | 0.01180629 |
| 1 | 219734960 | rs2820441 | C | A | 0.32 | 0.0861777 | 6.60E−13 | 0.01198998 |
| 2 | 56102744 | rs11899888 | G | A | 0.16 | 0.14842001 | 2.20E−12 | 0.02113889 |
| 2 | 43665943 | rs76684055 | G | A | 0.9 | 0.11332869 | 2.80E−10 | 0.01796169 |
| 3 | 55602137 | rs61613824 | A | T | 0.37 | 0.07696104 | 1.10E−10 | 0.01192727 |
| 3 | 100297679 | rs13083051 | T | G | 0.92 | 0.11332869 | 2.90E−08 | 0.02042927 |
| 4 | 4949339 | rs4330303 | G | A | 0.68 | 0.06765865 | 2.40E−08 | 0.01212439 |
| 4 | 174616174 | rs56063997 | C | T | 0.36 | 0.06765865 | 3.60E−10 | 0.01079007 |
| 5 | 64355060 | rs370763 | A | T | 0.67 | 0.09531018 | 3.30E−17 | 0.0112988 |
| 6 | 6743149 | rs1294421 | T | G | 0.4 | 0.06765865 | 5.60E−10 | 0.0109104 |
| 6 | 26099279 | rs13212652 | T | G | 0.87 | 0.11332869 | 3.10E−11 | 0.0170631 |
| 6 | 143676186 | rs6570555 | A | T | 0.43 | 0.07696104 | 7.80E−13 | 0.01074181 |
| 6 | 45481873 | rs62400367 | A | G | 0.85 | 0.0861777 | 2.90E−08 | 0.01553488 |
| 7 | 73540726 | rs3895707 | C | T | 0.91 | 0.10436002 | 2.30E−08 | 0.0186765 |
| 8 | 25435170 | rs10481336 | C | T | 0.21 | 0.09531018 | 1.60E−15 | 0.01196016 |
| 9 | 16766118 | rs7850168 | C | A | 0.08 | 0.11332869 | 1.70E−08 | 0.02009356 |
| 11 | 32350027 | rs7924571 | C | A | 0.78 | 0.07696104 | 8.80E−09 | 0.01337901 |
| 12 | 66328027 | rs12810758 | T | C | 0.23 | 0.07696104 | 3.20E−09 | 0.01299799 |
| 16 | 84856552 | rs4238714 | C | T | 0.42 | 0.0861777 | 2.80E−13 | 0.01179921 |
| 17 | 12191339 | rs12453693 | T | C | 0.31 | 0.07696104 | 3.00E−11 | 0.01157906 |

*Abbreviations: chr, chromosome; pos, position; eaf, effect allele frequency; se, standard error.

**S1 Table 2 Full results of three different Mendelian randomization methods.**

| Outcome | Method | beta | se | pval | or | or_lci95 | or_uci95 |
|---|---|---|---|---|---|---|---|
| AA | MR Egger | −0.3142796 | 0.50889807 | 0.54677418 | 0.73031484 | 0.26935643 | 1.98012638 |
| | Weighted median | 0.00861862 | 0.09606305 | 0.92851098 | 1.00865587 | 0.83555012 | 1.21762494 |
| | Inverse variance weighted | 0.05058115 | 0.11095215 | 0.6484744 | 1.05188222 | 0.84629689 | 1.30740905 |
| TAA | MR Egger | −0.3801559 | 0.49435936 | 0.45467937 | 0.68375484 | 0.25947363 | 1.80180421 |
| | Weighted median | 0.00623443 | 0.09477286 | 0.94755071 | 1.0062539 | 0.83567092 | 1.21165748 |
| | Inverse variance weighted | 0.04696191 | 0.10867452 | 0.66564435 | 1.04808208 | 0.84701222 | 1.29688336 |
| AAA | MR Egger | −0.1480964 | 0.54643699 | 0.79032927 | 0.86234802 | 0.2954922 | 2.51662857 |
| | Weighted median | 0.13508533 | 0.14021171 | 0.33532761 | 1.14463446 | 0.86959333 | 1.50666754 |
| | Inverse variance weighted | 0.14395554 | 0.11793032 | 0.22220604 | 1.15483277 | 0.91650489 | 1.45513541 |

*Abbreviations:MR, Mendelian randomization; AA, aortic aneurysm; TAA, thoracic aortic aneurysm; AAA, abdominal aortic aneurysm;or, Odds ratio; or_lci95, Lower limit of 95% confidence interval for OR; or_uci95, Upper limit of 95% confidence interval for OR.

**S1 Table 3 Pleiotropy test results.**

| Outcome | MR–Egger intercpt | SE | p |
|---|---|---|---|
| AA | 0.03177714 | 0.04322156 | 0.4743457 |
| TAA | 0.03719757 | 0.04198495 | 0.3905963 |
| AAA | 0.02542877 | 0.04640113 | 0.5923091 |

*p < 0.05 is set as the significant threshold. Pval > 0.05 represents no significant pleiotropy.

**S1 Table 4 Heterogeneity test results.**

| Outcome | Method | Q | p |
|---|---|---|---|
| AA | MR Egger | 46.24985 | 2.55E–05 |
| | Inverse variance weighted | 48.03556 | 2.51E–05 |
| TAA | MR Egger | 42.1749 | 1.16E–04 |
| | Inverse variance weighted | 44.53955 | 9.05E–05 |
| AAA | MR Egger | 22.64046 | 0.06637749 |
| | Inverse variance weighted | 23.12614 | 0.0814982 |

*p < 0.05 is set as the significant threshold. Pval > 0.05 represents no significant heterogeneity.

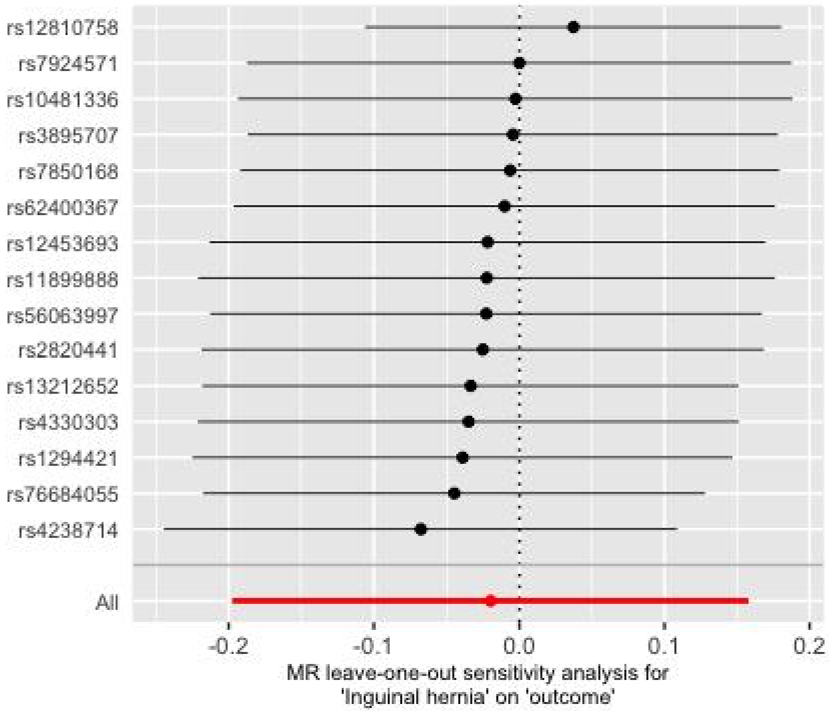


**S1 Figure 1 MR leave-one-out sensitivity analysis for the effect of the inguinal hernia SNPs on AA.**

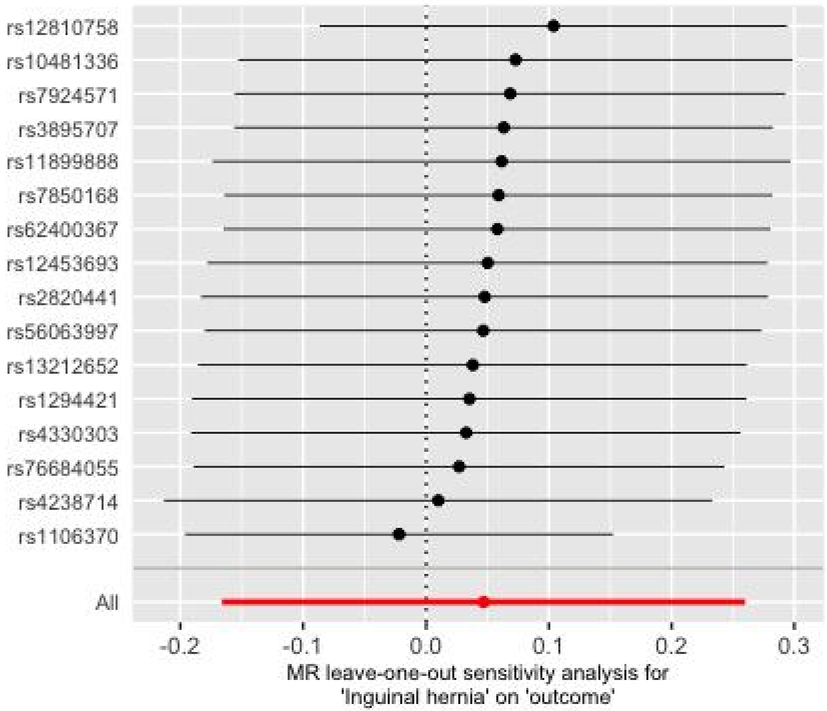


**S1 Figure 2 MR leave-one-out sensitivity analysis for the effect of the inguinal hernia SNPs on TAA.**

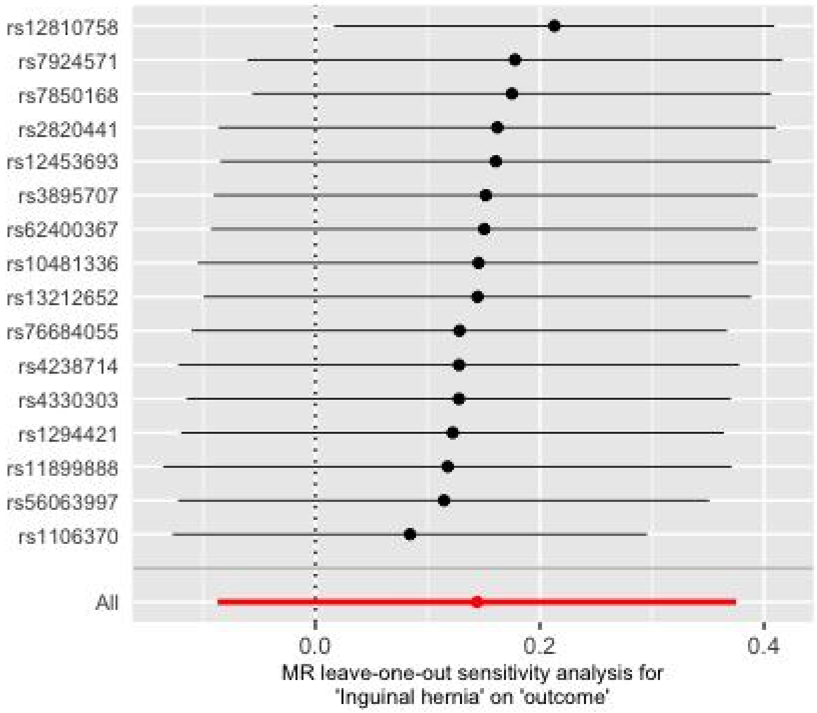


**S1 Figure 3 MR leave-one-out sensitivity analysis for the effect of the inguinal hernia SNPs on AAA.**
